# Level and duration of IgG and neutralizing antibodies to SARS-CoV-2 in children with symptomatic or asymptomatic SARS-CoV-2 infection

**DOI:** 10.1101/2022.04.12.22273466

**Authors:** Alka Khaitan, Dibyadyuti Datta, Caitlin Bond, Michael Goings, Katrina Co, Eliud O. Odhiambo, Lin Zhang, Stephanie Beasley, Josh Poorbaugh, Chandy C. John

## Abstract

**Background:** There are presently conflicting data about level and duration of antibodies to SARS-CoV-2 in children after symptomatic or asymptomatic infection.

**Methods:** We enrolled adults and children in a prospective 6-month study in the following categories: 1) symptomatic, SARS-CoV-2 PCR+ (SP+; children, n=8; adults, n=16), 2) symptomatic, PCR- or untested (children, n=27), 3) asymptomatic exposed (children, n=13) and 4) asymptomatic, no known exposure (children, n=19). Neutralizing and IgG antibodies to SARS-CoV-2 antigens and Spike protein variants were measured by multiplex serological assays.

**Results:** All SP+ children developed nAb, whereas 81% of SP+ adults developed nAb. Decline in the presence of nAb over 6 months was not significant in symptomatic children (100% to 87.5%, p=0.32) in contrast to adults (81.3 to 50.0%, p=0.03). Among all children with nAb (n=22), nAb titers and change in titers over 6 months were similar in symptomatic and asymptomatic children. Levels of IgG antibodies in children to the SARS-CoV-2 Spike, RBD-1 and -2, nucleocapsid and N-terminal domain antigens and to Spike protein variants were similar to those in adults. IgG levels to primary antigens decreased over time in both children and adults, but levels to three of six Spike variants decreased only in children.

**Conclusions:** Children with asymptomatic or symptomatic SARS-CoV-2 infection develop robust neutralizing antibodies that remain present longer than in adults but wane in titer over time, and broad IgG antibodies that also wane in level over time.

**Key Points:** Children have robust neutralizing and IgG antibody responses to SARS-CoV-2 infection after symptomatic or asymptomatic disease that are at least as strong as in adults. Neutralizing antibodies in children last longer than in adults but wane over time.

## Introduction

Since the onset of the COVID-19 pandemic, 28 million children had COVID-19 infection with nearly 13,000 pediatric deaths globally^1^. The latest COVID-19 surges increasingly affected children yet COVID-19 vaccines were only recently approved for children over the age of 5 years and no vaccine is available for children under 5. Understanding the breadth and durability of SARS-CoV-2 immunity in children after natural infection is critical to guide future protective measures for children.

Children typically have milder symptoms and less severe disease with SARS-CoV-2 infection than adults. One mechanism thought to mediate protection is the development of neutralizing antibodies (nAb) to SARS-CoV-2, which have been shown to be predictive of protection from symptomatic COVID-19 infection^2^, and are often used as a correlate of protective immunity to SARS-CoV-2. While there are numerous studies of humoral immunity in adults, studies in children are limited, with conflicting results^3-17^. Two recent and large studies in children demonstrated that children develop neutralizing antibodies at similar levels and with similar duration to those seen adults after infection^8,12^. Yet in earlier studies, children with severe or mild to severe COVID-19 had lower nAb levels and duration than adults^4,11,17^. It is not clear whether age, severity of COVID-19 infection, presence of specific SARS-CoV-2 variants or other factors are responsible for the different study findings.

To evaluate how level and duration of neutralizing and IgG antibodies to SARS-CoV-2 differ in children vs. adults, and whether symptomatic vs. asymptomatic infection affects this course in children, we conducted a study in adults and children starting early in the pandemic in the United States (June 2020) and evaluated these responses over a 6 month period of variable SARS-CoV-2 transmission.

## Materials and Methods

### Study population, enrollment and follow-up

From June 18 to December 29, 2020, children (<18 years old) and adults were enrolled in the Development of Immunity to SARS-CoV-2 after Exposure and Recovery (DISCOVER) study in Indianapolis, Indiana. Written informed consent was obtained from all participants and/or parents and verbal assent from children aged 7 to 17 years. Individuals were enrolled in 4 subject categories according to symptoms of and exposure to COVID-19 infection: 1) symptomatic PCR+ (SP+), 2) **symptomatic** PCR- or untested (SP-), 3) asymptomatic exposed (AE) and 4) asymptomatic, no known exposure(ANE). Symptomatic children included those who reported persistent cough, fever, chills, sore throat, shortness of breath, headache, new loss of taste or smell, muscle pain or diarrhea 21 days to 3 months prior to study enrollment. **SP+** had a documented positive SARS-CoV-2 naso- or oropharyngeal PCR test and **SP-** had a negative PCR test or were not tested. **Asymptomatic** children reported no symptoms of COVID-19 in the 3 months prior to study enrollment. **AE** children were in close contact with an individual with symptoms and a positive SARS-CoV-2 PCR. **ANE** children had no known close contact with an individual with confirmed or suspected COVID-19 requiring quarantine. Exclusion criteria included acute illness at study enrollment, immunocompromising conditions, or receipt of convalescent plasma.

Blood samples were collected at study enrollment and 6 months after enrollment. Vaccination was available for adults but not children during the later follow-up period, and date of vaccination for those vaccinated was recorded at the 6-month visit.

### Serological Assays

Luminex⍰xMAP⍰technology is a multiplex, flow cytometry-based platform that allows the simultaneous quantitation of many protein analytes in a single reaction^18^. A custom Luminex-based assay was developed to measure serology and antibody ACE2-RBD binding inhibition in a single assay as previously described^19,20^.

#### Surrogate neutralizing antibodies

Patient serum samples were titrated (1:20 – 1: 4.3E8) in phosphate buffered saline-high salt solution (PBS-HS; 0.01 M PBS, 1% [bovine serum albumin] BSA, 0.02% Tween, 300 mM NaCl). Diluted serum samples were combined with Luminex⍰MAGPlex⍰microspheres coupled with individual antigens and a recombinant, labelled RBD-Phycoerythrin (PE) protein and incubated for 60 minutes to allow endogenous antibodies to bind to either recombinant RBD-PE or antigen-coated Luminex beads. The solution was placed on a magnet, collecting the MAGPlex beads, while the supernatant was transferred to a new plate. The transferred solution was combined with ACE2 coated beads and incubated for 60 minutes; remaining beads were washed and incubated for 60 minutes with anti-IgG-PE beads to detect bound antibodies. All beads on both plates were washed and resuspended in a PBS-1% BSA solution and read using a Luminex⍰FlexMAP⍰3D System with xPONENT Software. The titer was evaluated from the median fluorescence intensity (MFI) and the ability of the endogenous antibodies to inhibit RBD-ACE2 binding was calculated based on the half maximal inhibitory concentration, IC50, which represents the antibody titer where the ACE2-RBD binding is reduced by half. The ACE2 binding inhibition potency was assessed using the inverse of IC50.

#### Immunoglobulin G to SARS-CoV-2 antigens

Antigen-coated microspheres were used to detect and quantitate endogenous IgG antibodies against SARS-CoV-2 proteins, including spike-NTD and several mutant RBD epitopes (Table S1). Testing was done before widespread Delta and Omicron variant infections and antibodies to these variants were not tested.

### Statistical Analysis

For the serial dilution-based serology assay, titer was calculated based on interpolating assay values that straddled the pre-determined “cut point”^19^. To calculate⍰IC50 of data from the ACE2 neutralization component of the serology assay, a 4-parameter logistic⍰function⍰was used to⍰estimate⍰the absolute IC50based on 1/dilution factor. If a sample indicated no neutralization, the IC50 was imputed to 1 (20 times the maximum 1/dilution factor).

Pairwise comparisons in the same individual were done using McNemar’s test for antibody prevalence and Wilcoxon matched-pairs signed-rank testing for titers. For comparisons across groups (children compared to adults, vaccine vs. no vaccine in adults), a Chi-square test or rank-sum was used for comparisons of prevalence or titers, respectively. No adjustment for multiple comparisons was done in the primary analysis because only two comparisons (by time point or by group at each time point) were performed, and none was made for the secondary analysis because it was exploratory. Evaluation of antibody decay/half-life was performed using exponential growth/decay equation for each individual in GraphPad Prism 9. Statistical analyses were conducted using Stata V16 (College Station, TX: StataCorp LP) and figures were created using GraphPad Prism 9.

## Results

### Demographic characteristics of children and adults in the study and SARS-CoV-2 epidemiology in Indiana at the time of the study

94 children and 344 adults were enrolled in the study (Figure 1). For this study on duration of antibodies, we included only children and adults who had samples collected at enrollment and 6 months after enrollment, and with the focus on children and limited testing capacity, we tested children from all 4 groups but only adults with COVID-19 (SP+). From the individuals with samples at both time points, all children with COVID-19 (SP+, n=8) and 23 of 58 SP+ adults were tested, as well as 27/29, 13/13 and 19/27 children from the SP-, AE and ANE groups (Figure 1). Local state and county case rates of COVID-19 during the study period relative to sample collection times are shown in Figure S1. Pediatric participants were 6 months to 17 years old (Table 1). ANE children were significantly older than AE children. There were no differences in sex between categories. 13% of adults and 13% of children were Black, and 13% of adults and 6% of children were Hispanic. There were no Asian or Native American participants in the group tested.

**Table 1.**
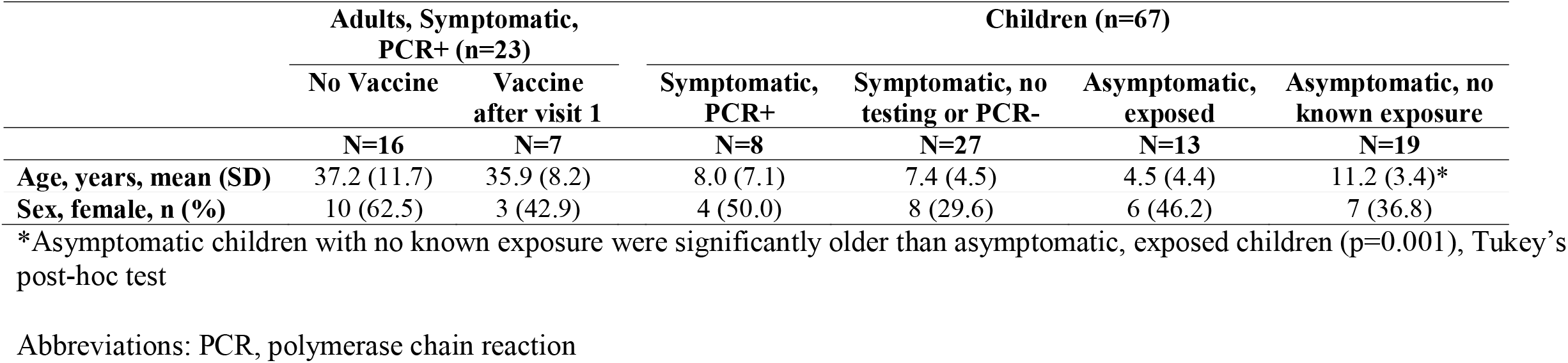
Demographic characteristics according to age and cohort

**Figure 1.**
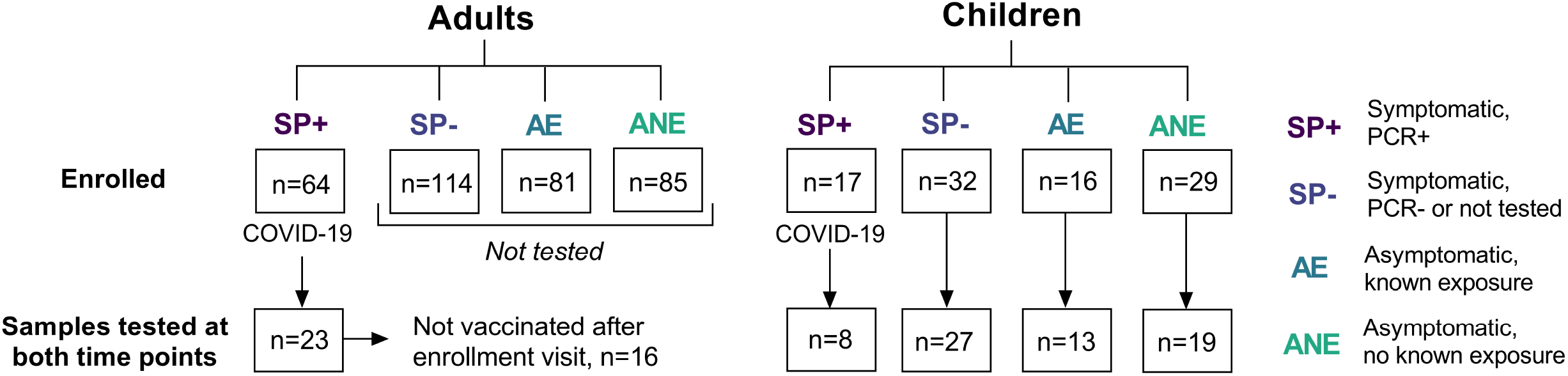
Study flowchart.

### Children with symptomatic SARS-CoV-2 infection develop neutralizing antibodies more frequently than asymptomatic exposed or non-exposed children, but titers among antibody-positive children do not differ in symptomatic vs. asymptomatic children

One hundred percent of SP+ children developed neutralizing antibodies (nAb) to SARS-CoV-2 compared to 30%, 39% and 5% in the SP-, AE and ANE groups, respectively (Fig 2A). Antibody titers among nAb positive symptomatic vs. asymptomatic children did not differ (Figure 2C).

**Figure 2.**
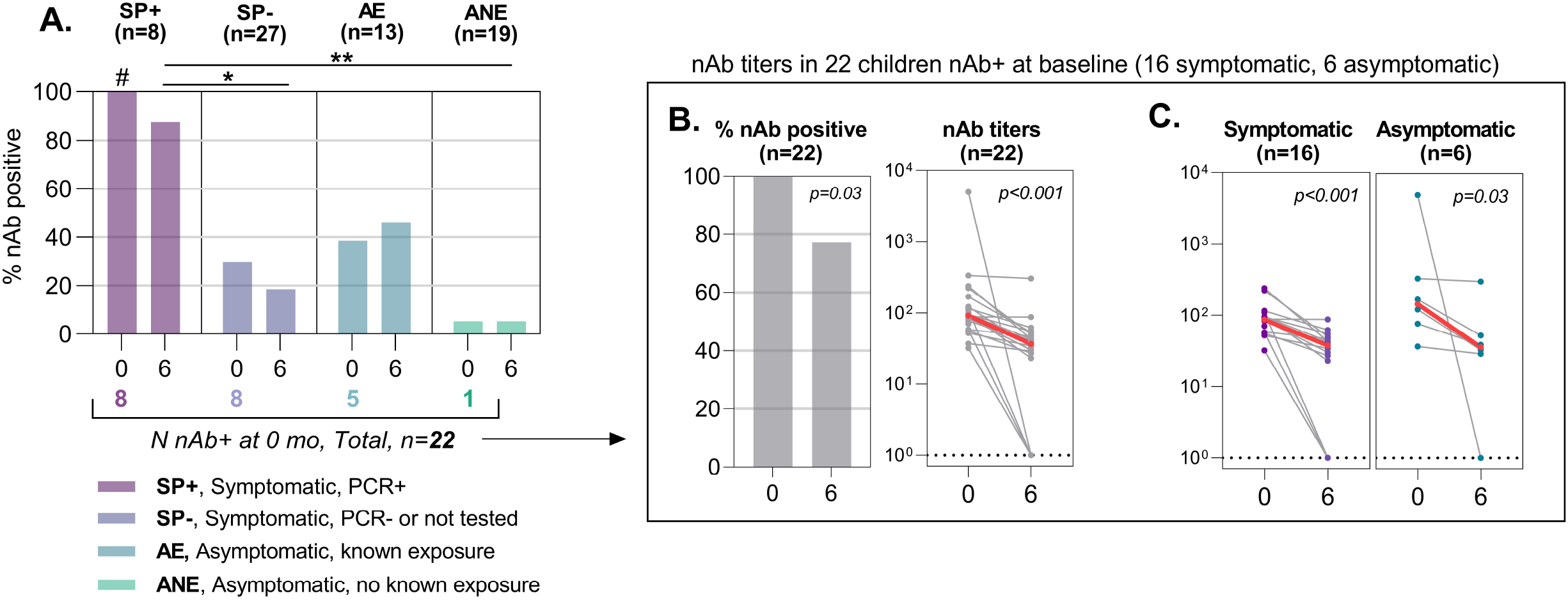
Neutralizing antibody (nAb) prevalence and titers at baseline and 6-month follow-up in (A) children according to study group, (B) all children with nAb at baseline, and (C) comparing children who were in the symptomatic groups to asymptomatic groups. Prevalence compared by visit using McNemar’s test and by study cohort using Fisher’s exact test, with Bonferroni correction for pairwise comparisons. Red dots and lines represent median of titers. Titers compared by visit using Wilcoxon matched-pairs signed-rank test and by study cohort using Wilcoxon rank-sum test. #significantly different than all other groups; *p<.05, **p<.001

### Neutralizing antibody seroprevalence and titers decrease in children with symptomatic or asymptomatic SARS-CoV-2 infection after 6 months

Among the 22 children with nAb at enrollment, 17 had nAb at 6 months (Figure 2B, p=0.03). Titers decreased significantly in children with symptomatic or asymptomatic infections (Figure 2C).

### Children with symptomatic SARS-CoV-2 infection develop neutralizing antibodies to SARS-CoV-2 at a similar proportion and level to adults, and for a longer duration

We next compared nAb presence titers and duration between SP+ children and adults (unvaccinated between visits, n=16). All children (100%) developed nAb whereas only 81% of adults developed nAb (Fig 3; Table S1). At 6 months, 88% of SP+ children had nAb and only 50% of SP+ adults had nAb present (Fig 3; Table S2). Thus a greater proportion of adults lost nAb by 6 months. Neutralizing antibody titers decreased in both SP+ children and adults over 6 months, and the mean difference in neutralizing antibody titers was greater in adults than children, but this difference was not statistically significant (Fig 3; Table S2).

**Figure 3.**
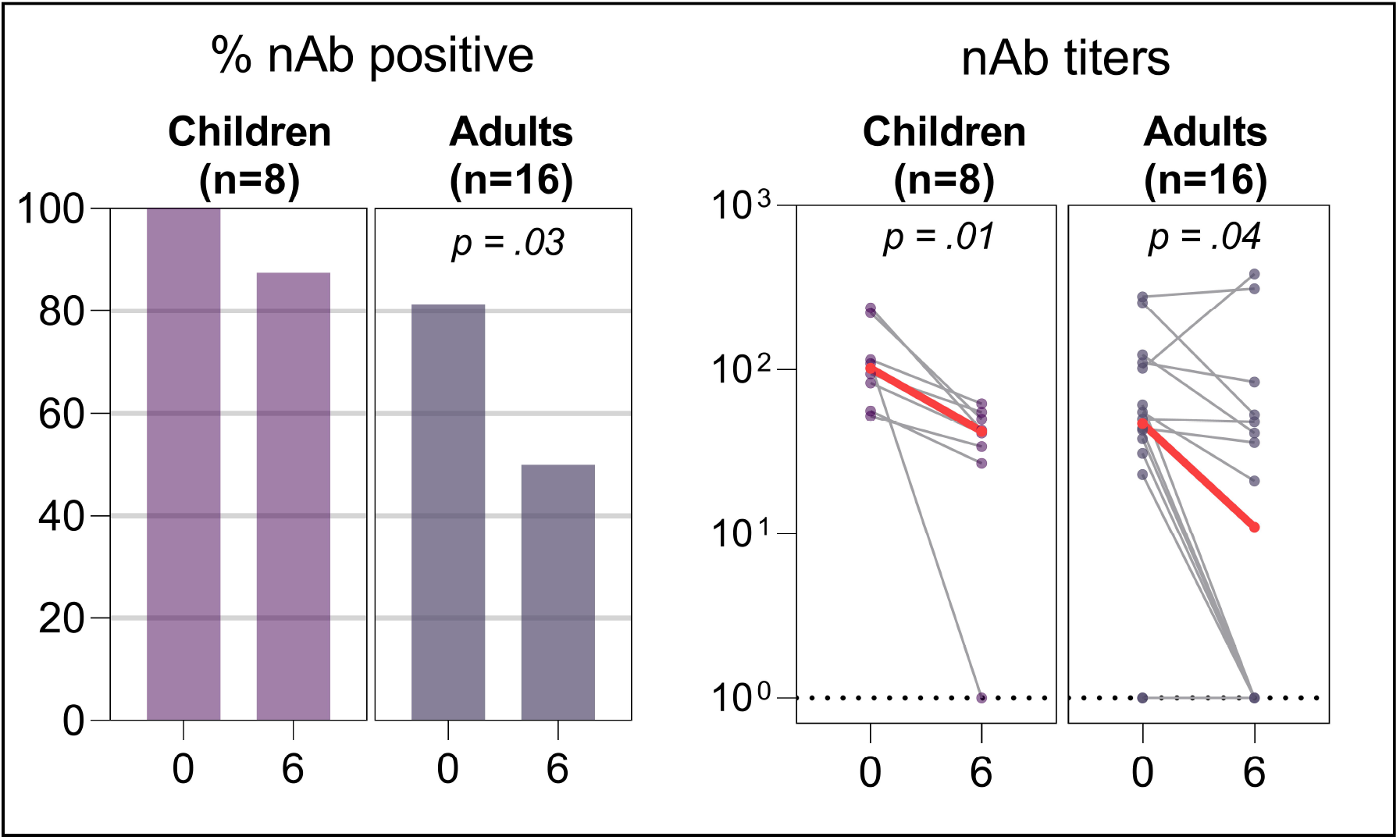
Prevalence and titers of neutralizing antibodies (nAb) at study baseline and 6-month follow-up in SARS-CoV-2 PCR-positive, symptomatic unvaccinated children and adults. Prevalence compared by visit using McNemar’s test and by age group using chi-squared test. Red dots and lines represent median of titers. Titers compared by visit using Wilcoxon matched-pairs signed-rank test and by age group using Wilcoxon rank-sum test.

### Children with COVID-19 develop IgG antibodies to SARS-CoV-2 antigens and variants at similar or higher levels to adults with COVID-19, and have similar decreases in antibody levels 6 months after initial infection

We next compared IgG antibodies to different SARS-CoV-2 proteins between adults and children with COVID-19 (the SP+ group), specifically examining total spike protein (ST4), RBD 1 and 2, nucleocapsid protein (NCP) and N-terminal Domain (NTD; Table S1). At initial visit, children had similar antibody levels as adults, and higher levels of RBD1 IgG compared with adults (Fig 4, Table S3). Children and adults had significant decreases in antibody levels to RBD2 and NCP, while only children had significant decreases RBD1, and there were no significant differences in antibody levels to ST4 or NTD in children or adults (Fig 4, Table S3). SP-, AE and ANE had no changes in these IgG levels in the 2^nd^ visit (Fig S1).

**Figure 4.**
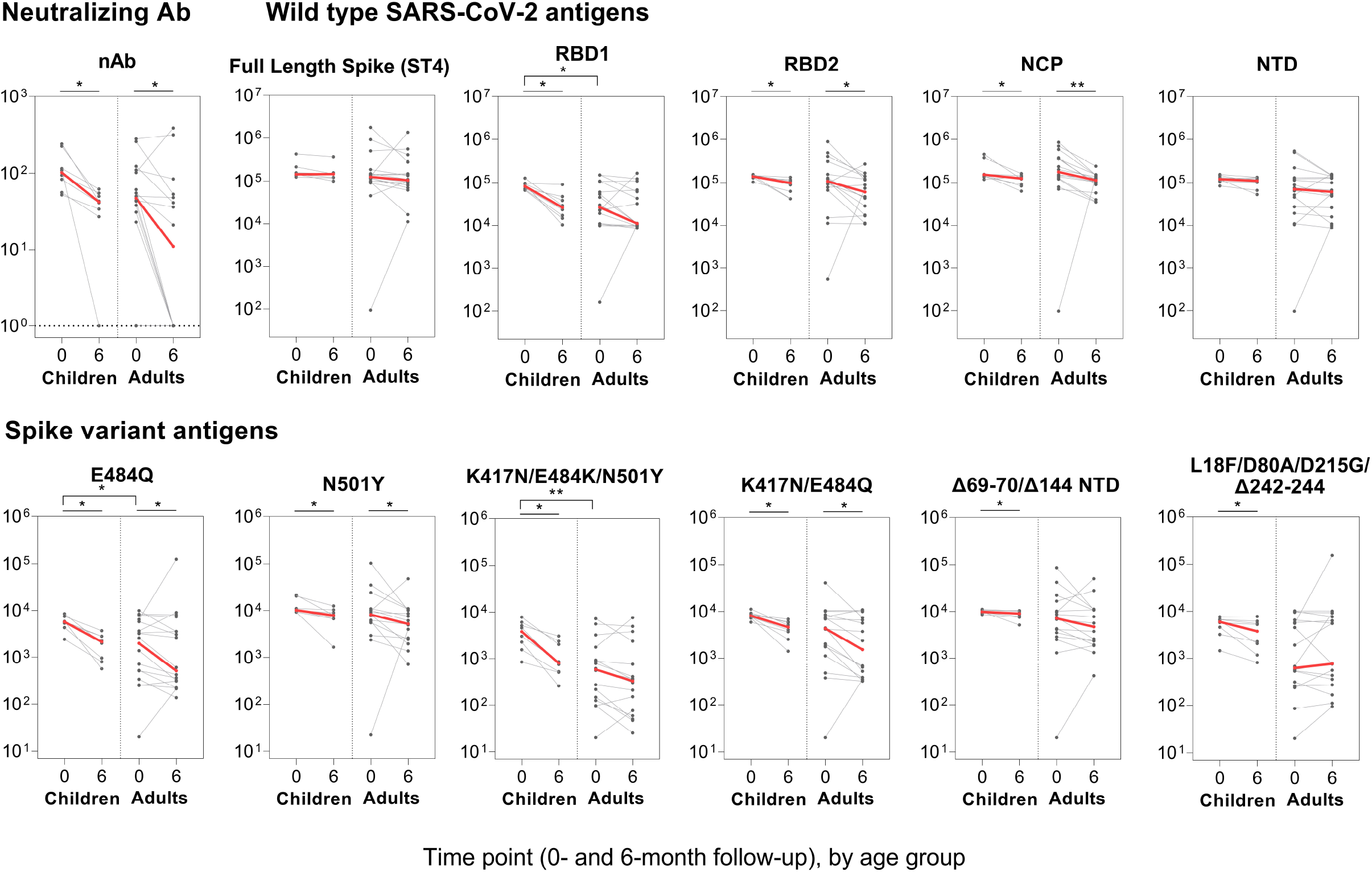
Titers at study baseline and 6-month follow-up in SARS-CoV-2 PCR-positive, symptomatic unvaccinated children (n=8) and adults (n=16). Red dots and lines represent median of titers. Titers compared by visit using Wilcoxon matched-pairs signed-rank test and by age group using Wilcoxon rank-sum test; *p<.05, **p<.01. Antigen definitions included in methods section

Given the continual emergence of novel SARS-CoV-2 variants, we also evaluated IgG antibodies to a several Spike protein variants (Table S1). These variants did not include the Delta or Omicron variants, which were not well established at the time of testing. SP+ children developed robust IgG antibodies to every variant tested, and had higher levels of antibodies to the E484Q and K417N variants than SP+ adults (Fig 4, Table S3). IgG antibodies to all Spike variants decreased significantly in SP+ children over time, but IgG antibodies to 3 of the Spike variants did not differ significantly over time in SP+ adults (Fig 4, Table S3). We also examined changes in IgG antibiotics in SP-, AE and ANE. SP-had decreases in levels of IgG to E484Q and d69-70 NTD, but asymptomatic children had no change in IgG levels at 6 months (Table S4), likely reflecting the large proportion of children who were neutralizing antibody negative in these groups at baseline (Fig S1). Neither neutralizing antibodies nor IgG antibodies in any child group correlated with age (all p>0.05)

### Vaccinated adults develop neutralizing and IgG antibody titers to multiple SARS-CoV-2 antigens higher than those after infection, and have a slow linear decrease in neutralizing antibody titers over time

Vaccines were unavailable to children during our study period, but 7 SP+ adults received COVID-19 vaccine between the enrollment and 6-month visits. We compared antibody titers at the 6-month visit between vaccinated and unvaccinated adults. All vaccinated adults had nAb at the 6 month visit whereas only 50% of unvaccinated adults had nAb in the follow up visit (Fig 5A). In vaccinated adults, nAb titers rose to levels higher than those with natural infection (Fig 5B). However, when we examined nAb titers relative to the time of vaccination, nAb titers decreased as the days post-vaccination increased (Fig 5C). Antibody responses to SARS-CoV-2 spike, RBD 1 and 2 and NTD increased after vaccination (Fig S3, Table S5), but antibodies to NCP decreased in both vaccinated and unvaccinated adults, as expected since the vaccine targets Spike protein. Antibody levels to Spike protein variants were also increased after vaccination, whereas unvaccinated adults had decreases in these antibody levels at the follow up visit (Fig S3, Table S5).

**Figure 5.**
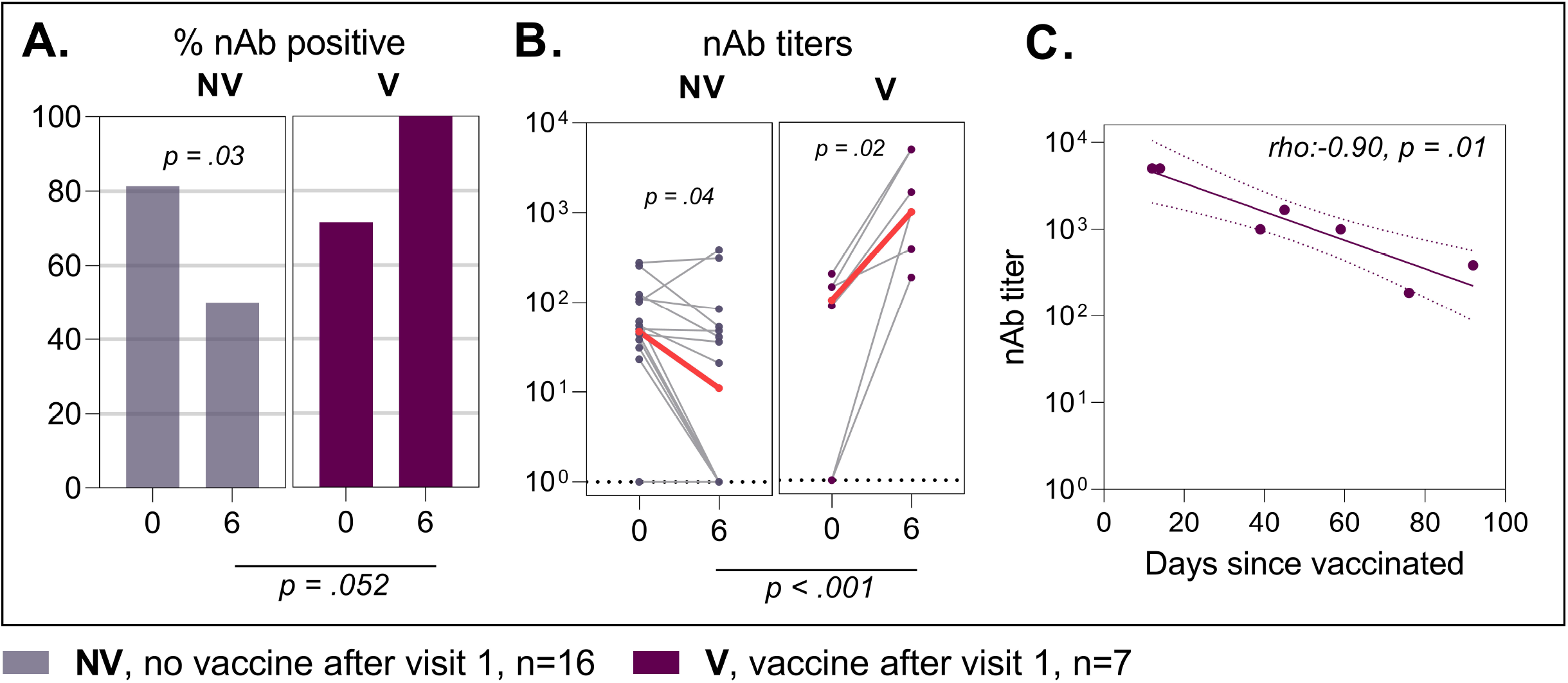
Prevalence (A) and titers (B) of neutralizing antibodies (nAb) at baseline and 6-month follow-up in adults who are not vaccinated (NV, n=16) and vaccinated after first visit (V, n=7), and (C) association between titers of nAb at 6-month follow-up and days since first vaccination dose in adults who were vaccinated. Prevalence compared by visit using McNemar’s test and by vaccination status using Fisher’s exact test. Red dots and lines represent median of titers. Titers compared by visit using Wilcoxon matched-pairs signed-rank test and by vaccination status using Wilcoxon rank-sum test. Mean (SD) days since first vaccine: 48 (30).

## Discussion

In this study, we show that children with symptomatic or asymptomatic SARS-CoV-2 infection develop robust neutralizing antibodies and that these neutralizing antibodies persist in children with symptomatic SARS-CoV-2 infection at a greater proportion than in adults at 6 months after infection, but titers wane over time in both children and adults. Children with symptomatic SARS-CoV-2 infection also develop IgG antibodies to multiple SARS-CoV-2 antigens and Spike antigen variants at similar or higher levels to adults, and with a generally similar decrease over 6 months, though for some antigens the decrease was greater in children than adults. Finally, we show that adults vaccinated against SARS-CoV-2 between the first and second visits develop levels of neutralizing and IgG antibodies significantly higher than with infection alone. The findings demonstrate the importance of evaluating neutralizing antibodies in studies of the immune response to SARS-CoV-2 in children and adults, as the IgG antibody levels to some antigens showed a greater decrease in children than adults, whereas a higher proportion of children than adults retained neutralizing antibodies to SARS-CoV-2 at 6 months.

Our results support recent studies showing similar or higher level and greater duration of neutralizing antibodies in children than adults after SARS-CoV-2 infection^8,9,12^, but differ from other studies that showed lower levels of neutralizing antibodies^17^ or IgG antibodies to the Spike protein^4^ and shorter duration of neutralizing or IgG antibodies in children compared to adults^4,11^. Our results suggest that children develop robust neutralizing antibody responses as well as antibodies to multiple antigens variants after initial SARS-CoV-2 infection. Importantly these responses remain present for at least 6 months, though titers decrease over time. Pediatric responses are at least as robust as adults, and a higher proportion of children maintain neutralizing antibodies at 6 months compared to adults. Children in this study were not vaccinated, so we could not compare titers after vaccine to titers after infection. As in other studies, adults developed much higher IgG and neutralizing antibody levels after infection followed by vaccination^21^. Pediatric vaccine studies also show significantly higher nAb levels after vaccination^22-24^, and demonstrate that vaccinated children have greater protection against severe COVID-19 than unvaccinated children^25^. Together, the study findings support a robust immune response in children, but show waning of neutralizing antibodies over time, supporting the need for vaccination and for studies on antibody kinetics in children after vaccination.

Potential reasons for differences in the pediatric studies may include the age of children, disease severity, various serological testing modalities and the predominant SARS-CoV-2 variant in circulation at the time of the study. Regarding the effects of age, in a subanalysis we found no correlations between age and levels of nAb or tested IgG. Our study showed, as did one prior study, a decrease in IgG antibodies to some SARS-CoV-2 antigens in children but not adults^4^, but the prior study did not evaluate neutralizing antibodies, which we show decreased less frequently in children than adults. We used a surrogate neutralizing assay here which was previously been reported to be strongly correlated with levels of neutralizing antibody measured with pseudovirus assays^20^. Two prior studies showed lower neutralizing antibody titers in children than adults with SARS-CoV-2 infection, one of which included only hospitalized children^11^ and the other of which included hospitalized children and children with asymptomatic infection^17^. In the second study, children also had a reduced breadth of antibodies, and predominantly generated antibodies to the S but not N antigen^17^. In contrast, the present study confirms the findings of three other studies that show children, whether symptomatic or asymptomatic, make neutralizing antibodies at similar or higher titers to adults, also generate antibodies to multiple SARS-CoV-2 antigens, and retain neutralizing antibodies longer than adults^8,9,12^. The present study adds to the prior studies in strongly supporting the robust acquisition of IgG and neutralizing antibodies in children after symptomatic or asymptomatic SARS-CoV-2 infection.

Children in our cohort developed antibodies to multiple SARS-CoV-2 variants, but we did not have the antigens for Delta or Omicron variants, so could not determine if IgG antibodies developed against these variants. High neutralizing antibody levels were shown to correlate with protection against these variants of concern (VOC), and children had relatively high neutralizing antibody titers, but it is unclear if they would be high enough to protect against VOC, particularly against Omicron. Recent data demonstrate that children develop nAb to Omicron, but at significantly lower levels than wild type^22,24^. Similarly, mRNA COVID-19 vaccines were shown to elicit neutralizing antibodies against Omicron in children and adolescents but at lower levels than nAb to wild type^22,23^. Based on the epidemiology of Omicron infections in children, which showed substantial transmission in areas in which many children had been previously exposed to virus^26,27^, it is unlikely that the neutralizing antibodies generated by non-Omicron variant infection would be sufficient to protect against Omicron.

The relatively small sample size (22 children with SARS-CoV-2 infection and samples at enrollment and follow-up), absence of children with severe SARS-CoV-2 infection, lack of testing for IgG against Delta or Omicron variants, and lack of testing of cellular immunity, which plays a critical role in memory responses to COVID-19 infection^8,28-31^, are all study limitations. However, the study strengths, notably the evaluation of antibodies over time in children and adults, evaluation of neutralizing antibodies and IgG antibodies against multiple Spike protein variants, and the inclusion of testing in asymptomatic children, provide valuable new data on level and duration of antibody response in children after SARS-CoV-2 infection.

In conclusion, our study shows that the neutralizing antibody response to SARS-CoV-2 infection in children after symptomatic or asymptomatic infection is at least as strong as in adults and lasts longer in children than in adults but does wane over time in both children and adults. IgG antibodies to some SARS-CoV-2 antigens showed a greater decrease in children than adults, but the more functionally important neutralizing antibodies showed a greater decrease in adults than children, highlighting the importance of evaluating neutralizing antibodies when assessing changes in immunity over time. Now that SARS-CoV-2 vaccines are available for children, studies of post-vaccination antibody responses in children will be important to determine the extent to which these vaccines augment and prolong the duration and breadth of antibody response, as they have in adults.

## Data Availability

All data produced in the present work are contained in the manuscript

## Acknowledgements

We thank the participants in this study. This work was supported by the Riley Children’s Foundation and the Indiana University School of Medicine Department of Pediatrics.

