## Supplemental Materials for "Level and duration of IgG and neutralizing antibodies to SARS-CoV-2 in children with symptomatic or asymptomatic SARS-CoV-2 infection"

### Supplemental Tables and Figures

**Table S1:** Details of SARS-CoV-2 proteins

| Protein | SARS-CoV-2 Sequence Length (AA) | Tags | Total Tag Length | Total Length | Expression |
|---|---|---|---|---|---|
| NTD | 294 (14-307) | AviTag, His Tag | 33 | 327 | CHO |
| NCP | 419 (1-419) | AviTag, His Tag | 33 | 452 | CHO |
| RBD-2 | 274 (319-592) | AviTag, His Tag | 33 | 307 | CHO |
| RBD-1 | 199 (329-527) | AviTag, His Tag | 33 | 232 | CHO |
| Full Length Spike (ST4) | 1195 (14-1208) | T4 trimerization, AviTag, His Tag | 62 | 1257 | CHO |
| E484Q | 274 (319-592) | AviTag, His Tag | 33 | 307 | CHO |
| UK variant (N501Y) | 274 (319-592) | AviTag, His Tag | 33 | 307 | CHO |
| K417N/E484Q | 274 (319-592) | AviTag, His Tag | 33 | 307 | CHO |
| South African variant (K417N/E484K/N501Y) | 274 (319-592) | AviTag, His Tag | 33 | 307 | CHO |
| (Δ69-70/Δ144) UK NTD | 291 (14-304) | AviTag, His Tag | 33 | 327 | CHO |
| South African variant (L18F/D80A/D215G/Δ242-244) | 291 (14-304) | AviTag, His Tag | 33 | 327 | CHO |

**Table S2.** Prevalence and titers of neutralizing antibodies (nAb) at study baseline and 6-month follow-up in SARS-CoV-2 positive, symptomatic unvaccinated children and adults.

| | **Children** | | | **Adults** | | |
|---|---|---|---|---|---|---|
| | **Enrollment** | **6 months** | P-value[a] | **Enrollment** | **6 months** | P-value[a] |
| **% nAb positive (n)** | 100 (8) | 87.5 (7) | 0.32 | 81.3 (13) | 50.0 (8) | 0.03 |
| **n Ab titers** | 102 (70, 169) | 42 (31, 53) | 0.01 | 47 (27, 106) | 11 (1, 51) | 0.04 |

[a] McNemar’s test to compare prevalence of nAb; Wilcoxon matched-pairs signed-rank test to compare nAb titers

**Table S3.** Titers at study baseline and 6-month follow-up in SARS-CoV-2 PCR-positive, symptomatic unvaccinated children (n=8) and adults (n=16).

| | Children | | | Adults | | |
|---|---|---|---|---|---|---|
| | **Enrollment** | **6 months** | | **Enrollment** | **6 months** | |
| | Median (IQR) | Median (IQR) | P-value[a] | Median (IQR) | Median (IQR) | P-value[a] |
| **Neutralizing antibody** | 102 (70, 169) | 42 (31, 53) | 0.01 | 47 (27, 106) | 11 (1, 51) | 0.04 |
| **Full Length Spike (ST4)** | 144033 (137270, 185275) | 146482 (119264, 152687) | 0.12 | 124611 (97707, 167777) | 105213 (74523, 213269) | 0.23 |
| **RBD1** | 81891 (70874, 94631) | 26190 (15961, 42711) | 0.01 | 26336 (10940, 83436)* | 11146 (9969, 64302) | 0.20 |
| **RBD2** | 137795 (129688, 148105) | 97882 (52345, 115923) | 0.01 | 106626 (71769, 232531) | 60237 (22711, 129895) | 0.03 |
| **NCP** | 152273 (140390, 264582) | 123773 (87479, 141530) | 0.03 | 176125 (122672, 361350) | 112547 (73287, 142380) | 0.003 |
| **NTD** | 119363 (107849, 137054) | 109900 (83530, 128239) | 0.09 | 69282 (23058, 124131) | 60562 (20735, 136334) | 0.16 |
| **E484Q** | 5708 (4828, 6674) | 2134 (880, 2795) | 0.01 | 1980 (426, 4710)* | 516 (267, 3560) | 0.03 |
| **N501Y** | 10214 (9634, 15763) | 7860 (6729, 9684) | 0.01 | 8196 (5721, 10486) | 5203 (2513, 9395) | 0.03 |
| **K417N/E484K/N501Y** | 3795 (1901, 5591) | 801 (516, 2191) | 0.01 | 584 (136, 1342)** | 330 (68, 1565) | 0.06 |
| **K417N E484Q** | 8099 (7402, 8895) | 4583 (3111, 5879) | 0.01 | 4310 (1429, 8049) | 1524 (388, 6947) | 0.02 |
| **Δ69-70/Δ144 NTD** | 9773 (9270, 10514) | 8981 (7658, 10095) | 0.01 | 7101 (3184, 13363) | 4696 (2134, 10782) | 0.12 |
| **L18F/D80A/D215G/Δ242-244** | 5985 (3910, 6690) | 3770 (1628, 5838) | 0.02 | 628 (286, 6198) | 771 (316, 7021) | 0.88 |

[a] Wilcoxon matched-pairs signed-rank test.

*p<.05, **p<.01, Wilcoxon rank-sum test to compare adults vs. children at enrollment.

**Table S4.** Titers at study baseline and 6-month follow-up according to study cohort.

| | Cohort | Enrollment | 6 mo. | |
|---|---|---|---|---|
| | | Median (IQR) | Median (IQR) | P[a] |
| **Neutralizing antibody** | SP+ | 102 (70, 169) | 42 (31, 53) | **0.01** |
| | SP- | 1 (1, 32) | 1 (1, 1) | **0.005** |
| | AE | 1 (1, 77) | 1 (1, 33) | 0.08 |
| | ANE | 1 (1, 1) | 1 (1, 1) | 0.32 |
| **Full Length Spike (ST4)** | SP+ | 144033 (137270, 185275) | 146482 (119264, 152687) | 0.12 |
| | SP- | 2872 (273, 133838) | 2903 (236, 63274) | 0.41 |
| | AE | 3496 (307, 154355) | 10044 (239, 123861) | 0.10 |
| | ANE | 669 (317, 1825) | 554 (304, 1250) | 0.63 |
| **RBD1** | SP+ | 81891 (70874, 94631) | 26190 (15961, 42711) | **0.01** |
| | SP- | 305 (162, 48222) | 247 (99, 9919) | 0.14 |
| | AE | 217 (77, 93334) | 6740 (87, 21665) | 0.35 |
| | ANE | 305 (118, 342) | 234 (146, 542) | 0.18 |
| **RBD2** | SP+ | 137795 (129688, 148105) | 97882 (52345, 115923) | **0.01** |
| | SP- | 765 (354, 85065) | 725 (363, 21926) | 0.28 |
| | AE | 1432 (328, 145097) | 8854 (649, 100402) | 0.25 |
| | ANE | 629 (376, 2171) | 599 (358, 2053) | 0.63 |
| **NCP** | SP+ | 152273 (140390, 264582) | 123773 (87479, 141530) | **0.03** |
| | SP- | 2577 (380, 133535) | 760 (342, 58721) | 0.13 |
| | AE | 6085 (334, 151857) | 10500 (355, 97042) | 0.13 |
| | ANE | 671 (270, 5285) | 579 (320, 4751) | 0.38 |
| **NTD** | SP+ | 119363 (107849, 137054) | 109900 (83530, 128239) | 0.09 |
| | SP- | 128 (73, 46482) | 90 (62, 10701) | 0.08 |
| | AE | 279 (22, 136466) | 3129 (28, 93801) | 0.94 |
| | ANE | 72 (37, 119) | 57 (30, 94) | 0.28 |
| **E484Q** | SP+ | 5708 (4828, 6674) | 2134 (880, 2795) | **0.01** |
| | SP- | 20 (20, 1242) | 20 (20, 312) | **0.04** |
| | AE | 25 (20, 5739) | 38 (20, 1903) | 0.13 |
| | ANE | 20 (20, 20) | 20 (20, 20) | 0.95 |
| **N501Y** | SP+ | 10214 (9634, 15763) | 7860 (6729, 9684) | **0.01** |
| | SP- | 180 (29, 9831) | 82 (21, 6649) | 0.05 |
| | AE | 77 (36, 10362) | 425 (104, 8473) | 0.86 |
| | ANE | 141 (34, 331) | 114 (38, 257) | 0.16 |
| **K417N/E484K/N501Y** | SP+ | 3795 (1901, 5591) | 801 (516, 2191) | **0.01** |
| | SP- | 24 (20, 437) | 20 (20, 329) | **0.001** |
| | AE | 21 (20, 2066) | 20 (20, 607) | **0.01** |
| | ANE | 20 (20, 20) | 20 (20, 20) | 0.32 |
| **K417N E484Q** | SP+ | 8099 (7402, 8895) | 4583 (3111, 5879) | **0.01** |
| | SP- | 31 (20, 4726) | 28 (20, 702) | **0.04** |
| | AE | 31 (20, 7986) | 101 (20, 5376) | 0.19 |
| | ANE | 20 (20, 21) | 20 (20, 31) | 0.94 |
| **Δ69-70/Δ144 NTD** | SP+ | 9773 (9270, 10514) | 8981 (7658, 10095) | **0.01** |
| | SP- | 41 (20, 6653) | 33 (20, 3560) | **0.02** |
| | AE | 39 (20, 10209) | 69 (20, 8874) | 0.83 |
| | ANE | 20 (20, 27) | 20 (20, 20) | 0.94 |
| **L18F/D80A/D215G/Δ242-244** | SP+ | 5985 (3910, 6690) | 3770 (1628, 5838) | **0.02** |
| | SP- | 39 (20, 1032) | 42 (20, 299) | 0.06 |
| | AE | 51 (20, 4863) | 46 (20, 3562) | 0.56 |
| | ANE | 20 (20, 20) | 20 (20, 23) | 0.31 |

[a] Wilcoxon matched-pairs signed-rank test

SP+, symptomatic, PCR+ (n=8); SP-, symptomatic , PCR- or not tested (n=27); AE, asymptomatic, known exposure (n=13); ANE, asymptomatic, no known exposure (n=19)

**Table S5.** Titers at study baseline and 6-month follow-up in SARS-CoV-2 PCR-positive, symptomatic adults who were unvaccinated (n=16) and vaccinated (n=7).

| | No vaccination after visit 1 (n=16) | | | Vaccinated after visit 1 (n=7) | | |
|---|---|---|---|---|---|---|
| | **Enrollment** | **6 months** | | **Enrollment** | **6 months** | |
| | Median (IQR) | Median (IQR) | P[a] | Median (IQR) | Median (IQR) | P[a] |
| **Neutralizing antibody** | 47 (27, 106) | 11 (1, 51) | 0.04 | 103 (1, 145) | 1000 (385, 5000) † | 0.02 |
| **Full Length Spike (ST4)** | 124611 (97707, 167777) | 105213 (74523, 213269) | 0.23 | 147063 (21665, 204154) | 2013954 (1348154, 14422662) † | 0.02 |
| **RBD1** | 26336 (10940, 83436) | 11146 (9969, 64302) | 0.20 | 79560 (716, 121739) | 756835 (153278, 1528487) † | 0.02 |
| **RBD2** | 106626 (71769, 232531) | 60237 (22711, 129895) | 0.03 | 145817 (378, 151965) | 1432052 (763876, 2074017) † | 0.02 |
| **NCP** | 176125 (122672, 361350) | 112547 (73287, 142380) | 0.003 | 141966 (470, 216876) | 57193 (318, 152774) | 0.02 |
| **NTD** | 69282 (23058, 124131) | 60562 (20735, 136334) | 0.16 | 90059 (200, 133184) | 1129327 (690146, 2086326) † | 0.02 |
| **E484Q** | 1980 (426, 4710) | 516 (267, 3560) | 0.03 | 5293 (27, 6435) | 34278 (10781, 123962) † | 0.02 |
| **N501Y** | 8196 (5721, 10486) | 5203 (2513, 9395) | 0.03 | 10723 (85, 18671) | 138390 (33938, 193936) † | 0.02 |
| **K417N/E484K/N501Y** | 584 (136, 1342) | 330 (68, 1565) | 0.06 | 1913 (327, 5033) | 18306 (8513, 123734) † | 0.02 |
| **K417N E484Q** | 4310 (1429, 8049) | 1524 (388, 6947) | 0.02 | 8780 (20, 10401) | 74318 (18926, 147037) † | 0.02 |
| **Δ69-70/Δ144 NTD** | 7101 (3184, 13363) | 4696 (2134, 10782) | 0.12 | 8607 (82, 10133) | 129870 (95337, 838793) † | 0.02 |
| **L18F/D80A/D215G/Δ242-244** | 628 (286, 6198) | 771 (316, 7021) | 0.88 | 1697 (66, 5554) | 67231 (20100, 154028) † | 0.02 |

[a] Wilcoxon matched-pairs signed-rank test.

†p<.001, Wilcoxon rank-sum test to compare adults with no vaccination vs. adults with vaccination at 6-month follow-up.

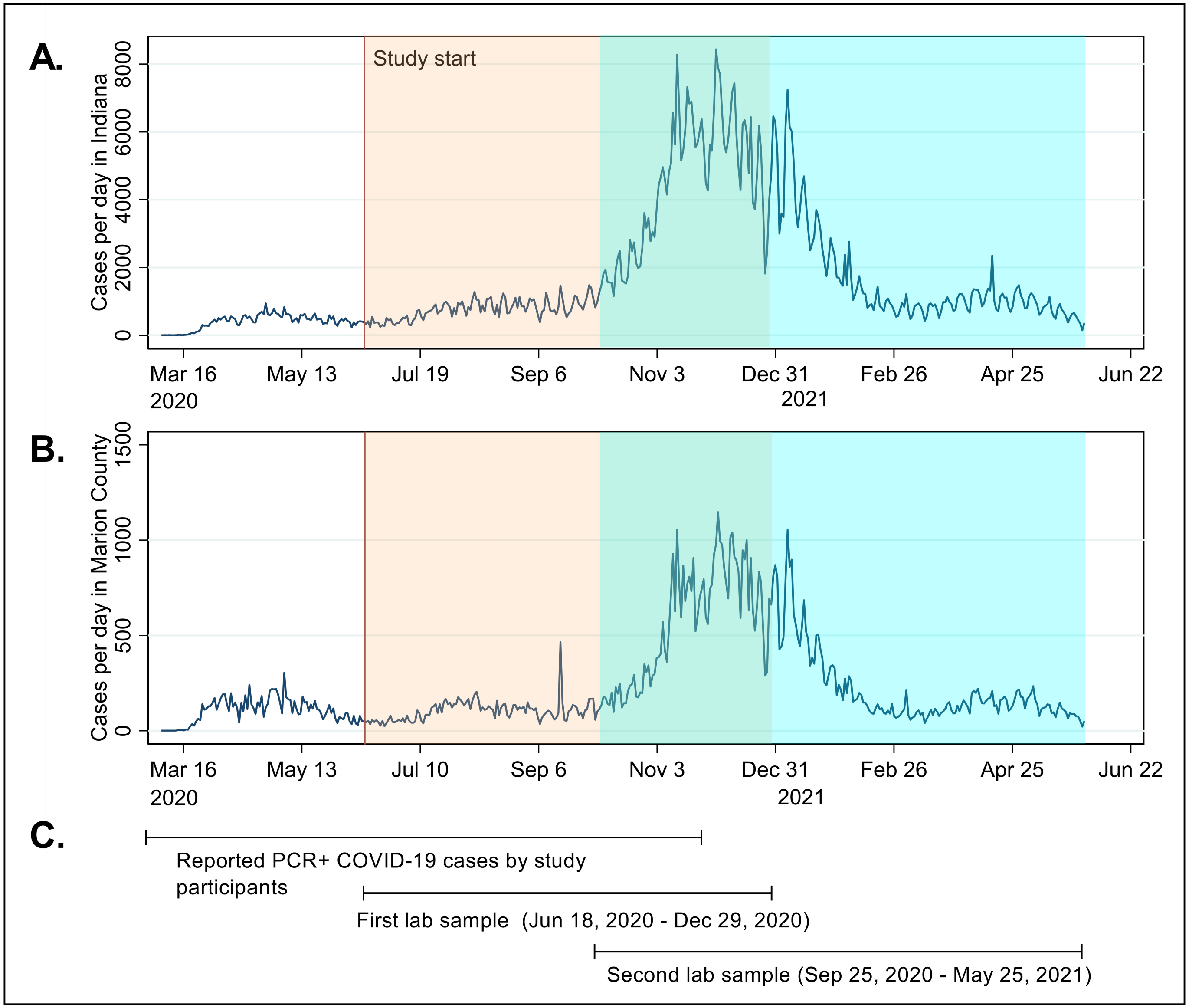


**Figure S1. Incidence of COVID-19 in Indiana during the study period**

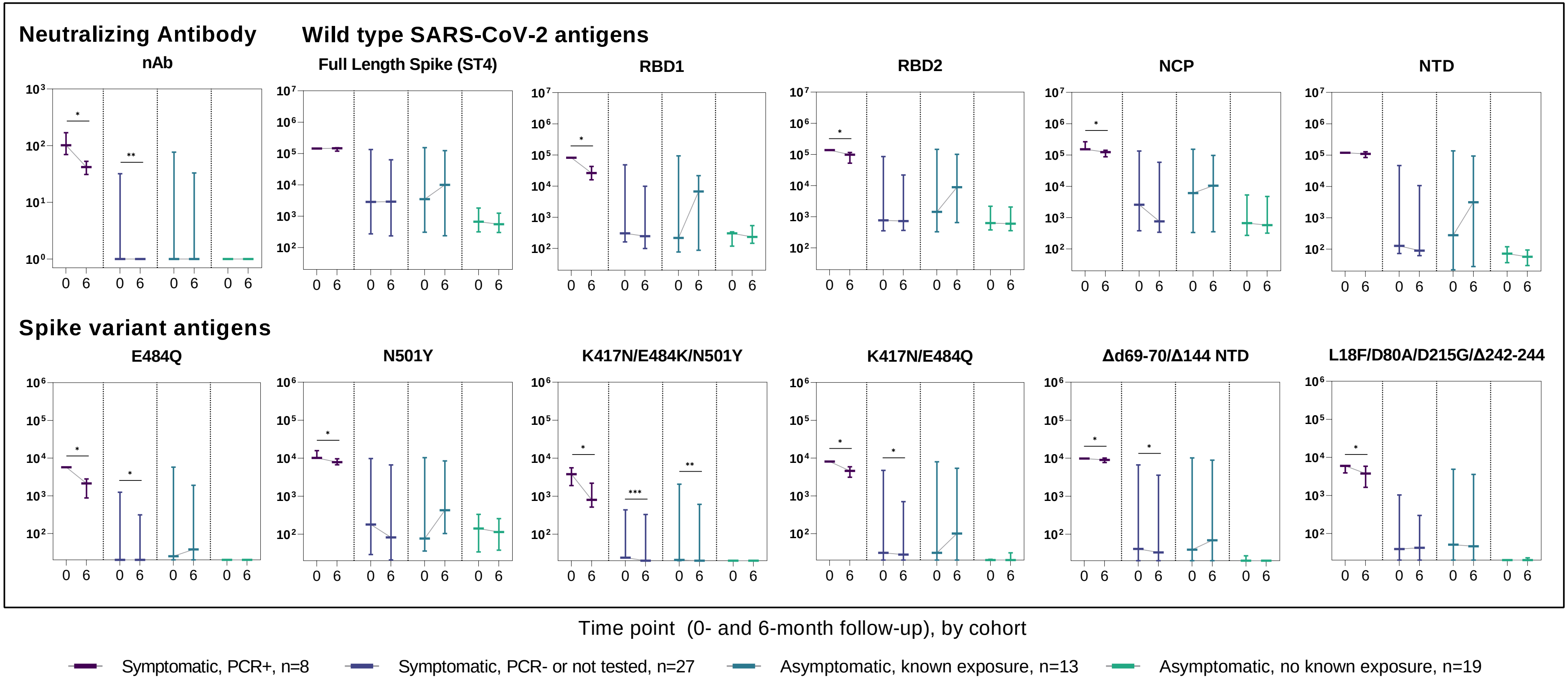


**Figure S2. Titer levels at study baseline and 6-month follow-up in children, according to study cohort. Line and error bars represent median and interquartile range.** Titers compared by visit using Wilcoxon matched-pairs signed-rank test; *p<.05, ** p<.01, *** p<.001.

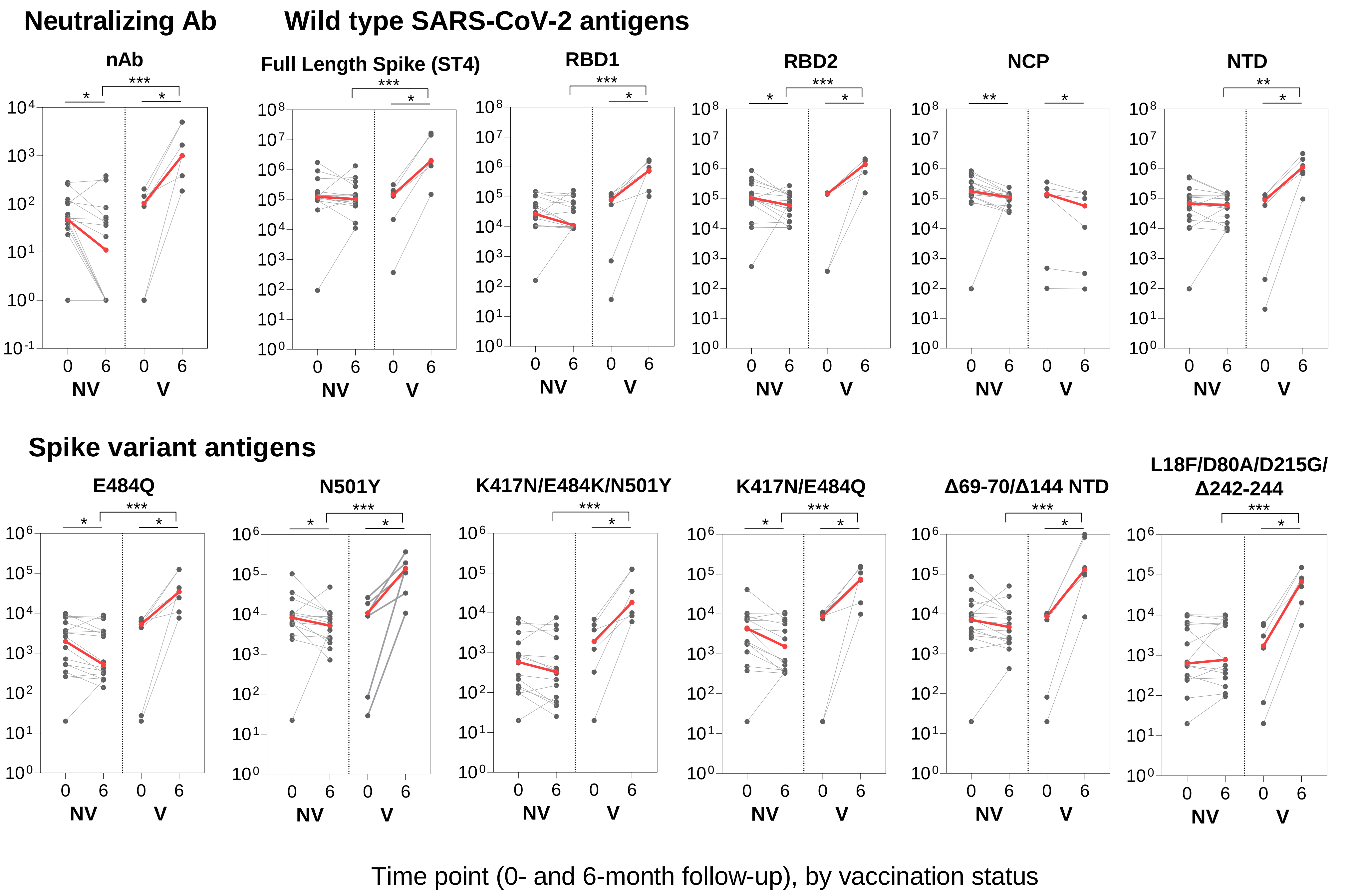


**Figure S3. Titers at study baseline and 6-month follow-up in SARS-CoV-2 PCR-positive, symptomatic adults with no vaccination after visit 1 (NV, n=16) and those vaccinated after visit 1 (V, n=7).** Red dots and line represent median of titers. Titers compared by visit using Wilcoxon matched-pairs signed-rank test and by age group using Wilcoxon rank-sum test; *p<.05, **p<.01. Antigen definitions included in methods section.
